# Understanding the Barriers and Enablers of Pharmacogenetics Testing in Primary Care in Singapore: A Qualitative Study

**DOI:** 10.64898/2025.12.07.25341810

**Authors:** Amarasinghe Arachchige Don Nalin Samandika Saparamadu, Oshozimhede E. Iyalomhe, Xin Hui Sam, Aloysius Chow, Shirley Sun, Helen Elizabeth Smith

**Affiliations:** Department of International Health, Johns Hopkins Bloomberg School of Public Health, Baltimore, Maryland, United States; Department of Nutrition and Food Studies, College of Public Health, George Mason University, Fairfax, Virginia, United States; University of Maryland Institute for Health Computing, North Bethesda, Maryland, United States; Department of Epidemiology and Human Genetics, University of Maryland School of Medicine, Baltimore, Maryland, United States; Primary Care and Family Medicine, Lee Kong Chian School of Medicine, Nanyang Technological University, Singapore; School of Social Sciences, Nanyang Technological University, Singapore; Lee Kong Chian School of Medicine, Nanyang Technological University, Singapore; School of Medicine, Keele University, Keele, Staffordshire, ST5 5BG, United Kingdom

**Keywords:** pharmacogenetics, pharmacogenomics, precision medicine, primary care, Singapore, qualitative research, drug utilization

## Abstract

Pharmacogenetic testing promises safer, personalized prescribing, yet its adoption in primary care in Singapore remains limited and underexplored. The aim of this study was to, 1) examine perceptions, attitudes, and beliefs about pharmacogenetic testing among primary care physicians (PCPs) and patients in Singapore, and 2) to compare acceptability between those who did and did not participate in a prior pharmacogenetic feasibility study.

We conducted one-to-one semi-structured interviews with a purposive sample of 10 PCPs (6 had joined an earlier feasibility study) and 15 patients (7 previously tested with pharmacogenetic testing). Interviews (mean=35 minutes) were audio-recorded, transcribed verbatim, and analysed thematically by three investigators. Ethical approval was obtained (IRB2019-07-035).

Five principal themes emerged: 1) Perceptions of acceptability and test accuracy—broad support for pharmacogenetics to reduce trial-and-error prescribing but persistent concerns about result reliability and medico-legal risk; 2) Appropriate financial context—cost sensitivity in Singapore with expectations for government subsidies and incentives for PCPs; 3) Physician education and communication—variable pharmacogenetic expertise, perceived inadequacy of training, need for expert support, and challenges explaining probabilistic results to patients; 4) Integration into clinical records—frustration with standalone pharmacogenetic software and a desire for EMR integration; 5) Data privacy and insurance implications—concerns about insurer access and potential premium and eligibility impacts. No significant attitude differences were observed between participating and non-participating PCPs.

Patients and PCPs generally favor pharmacogenetic testing in primary care but identify actionable barriers, cost, training, and data integration and safeguards, that must be addressed to support implementation in Singapore.

## Introduction

Pharmacogenetics involves using a patient’s genetic information to guide the selection of medications and is set to play a transformative role in medical practice by promoting more personalized and precise drug therapy (1,2). In 2016, the prevalence of adverse drug reactions (ADRs) in Singapore was 12.4% at the time of hospital admission (3). A subsequent study using 1000 randomly selected hospitalizations showed ADR-related admissions cost USD 570,404, or 10.1% of total expenses (4). Extrapolated, the annual ADR burden in Singapore is estimated at USD 168 million, representing nearly 5% of healthcare expenditure (5). Pharmacogenetic testing can predict a person’s therapeutic response by identifying variations in one or more genes that influence drug response (6). Therefore, pre-emptive pharmacogenetic testing, particularly in primary care settings where most medications are initiated, has the potential to reduce ADRs (4,7). However, despite the demonstrated feasibility of pharmacogenetic testing in primary care from other countries (8) and more recently in Singapore itself (9), its application in the community setting remains uncommon.

Healthcare providers in Western countries recognize the value of pharmacogenetics in optimizing drug prescriptions (10–12), and randomized controlled trials have demonstrated that pharmacogenetics-guided therapy can be successfully implemented in primary care and community health care settings (8), and when prescribing for people with psychiatric disorders (13,14). Nonetheless, despite the robust evidence emerging, implementation has been limited. The barriers contributing to this include providers’ knowledge and awareness, insufficient training, perceived negative impacts, cost-effectiveness concerns, and ethical issues (15,16).

An exploratory study of pharmacogenetics (9) in Singapore has confirmed the feasibility of pharmacogenetic testing in primary care. This qualitative study seeks to inform the implementation of pharmacogenetic testing in primary care by exploring the perceptions, attitudes, and beliefs of patients and primary care physicians (PCPs). It also examines differences in acceptability between those who had and had not participated in the earlier pharmacogenetic feasibility study.

## Methods

### Study design

In semi-structured interviews using a topic guide we sought patients’ and PCPs’ perceptions, attitudes, experiences and beliefs about pharmacogenetic testing.

### Study participants

A purposive sample of PCPs was recruited through the Primary Care Research Network and included some that had and some that had not participated in the forementioned pharmacogenetics feasibility study (9). Patients taking long-term medications for one or more chronic conditions were invited by their family doctors and recruited through the enrolled PCPs in the pcRn considering two factors: participation in the pharmacogenetics feasibility study and convenience.

### Data collection

One-to-one semi-structured interviews were conducted in person or using the video-conference platform Zoom. All PCPs were interviewed by the researcher SS, and patients were interviewed by the researchers SHX or SS. Both interviewers had >3 years’ experience in conducting qualitative research. SS is medically qualified, and SHX has postgraduate training in molecular biotechnology. Any patient who had prior clinical contact with SS was interviewed by SHX to avoid interviewer bias. All participants were informed about the research aims and the use of data. Written consent and demographic details were obtained before the interviews.

The interview guide, developed by the research team, explored the perception, acceptance, and experiences of participants with regards to pharmacogenetic testing, for PCPs as providers and patients as users in primary health care setting (Supplementary file 1-4). Interviews were conducted between June 2021 and June 2022. There was a briefing about pharmacogenetic testing before the interview began for those patients who had no prior experience of pharmacogenetic testing (n=8).

### Thematic analysis

The audio recordings were anonymized, transcribed verbatim and transcripts compared with the recordings to confirm accuracy. Three researchers, SHX, SS, and OEI, read and re-read the transcripts to become familiar with the content before coding independently. Afterwards, they worked collaboratively with the other members of the research team to interpret the data and finalize the emerging themes.

### Ethics approval

Institutional review board approval was granted by Nanyang Technological University Ethics Committee (reference IRB2019-07-035).

## Results

Ten primary care clinicians were interviewed, of whom six (60%) had participated in the pharmacogenetics feasibility study. Seven (47%) of the 15 patients with chronic conditions receiving long-term care within primary healthcare had previously undergone pharmacogenetic testing. (Table 1). Interviews lasted up to 52 minutes (mean=35).

**Table 1:** Demographic characteristics of the study participants – participating and non-participating primary care provider and patients from Singapore. **1.1 Overall participant characteristics**

| ***All participants (N=25)*** | ***N (%)*** |
| --- | --- |
| Gender |  |
| - Male | 16 (64) |
| - Female | 9 (36) |
| Role |  |
| - Service provider (General practitioner) | 10 (40%) |
| - Service recipient (Patient) | 15 (60%) |
| Duration |  |
| - Minimum | 16 minutes |
| - Maximum | 50 minutes |
| - Mean | 34.98 (SD: 8.39) |
| ***Primary care providers (N=10)*** |  |
| Gender |  |
| - Male | 9 (90) |
| - Female | 1 (10) |
| Practice setting |  |
| - Public | 2 (20) |
| - Private | 8 (80) |
| Pharmacogenetic (PGx) exposure/experience |  |
| - Prior participation in PGx testing | 6 (60) |
| - No prior experience with PGx testing | 4 (40) |
| Years of practice |  |
| - < 10 years | 1 (10) |
| - ≥ 10 years | 9 (90) |
| ***Primary care patients (N=15)*** |  |
| Gender |  |
| - Male | 7 (47) |
| - Female | 8 (53) |
| Age (years) |  |
| - 41-50 | 4 (27) |
| - 51-60 | 4 (27) |
| - 61-70 | 3 (20) |
| - 71-80 | 2 (13) |
| - Missing data | 2 (13) |
| Education level |  |
| - Secondary school | 4 (27) |
| - Diploma | 3 (20) |
| - Degree/Postgraduate | 6 (40) |
| - Missing data | 2 (13) |
| Ethnic |  |
| - Chinese | 9 (60) |
| - Malay | 2 (13) |
| Indian | 4 (27) |
| PGx exposure/experience |  |
| - Prior participation in PGx testing | 7 (47) |
| - No prior experience with PGx testing | 8 (53) |

**Table 2:** Selected quotes from participating and non-participating primary care providers and patients distributed across the five thematic areas.

| Thematic Areas | Selected Quotes |
| --- | --- |
| 1. Perceptions of acceptability and test accuracy | We treat patients in a trial-and-error kind of way, [...] pharmacogenetics reduces the amount of trial and error that you can have [chuckles]." [GN19] (participating PCP) |
|  | "...we are also moving towards precision medicine, and I think such tests will be one way of delivering person-centered care. So, if you know that you have these genes, you are more susceptible to this particular medication, for example, and this can be identified by the pharmacogenetic tests. And if it is affordable, [...] not difficult to be carried out, if it doesn't cause much inconvenience, [...] we are all out to support such testing." [GT05] (non-participating PCP) |
|  | "...based on the drugs available in the market and [...] the diagnosis that [...] the doctor has put in the system can, by using the AI. ...going forward, [...] the system can detect and tell what is the best medications" [PT07] (non-participating patient) |
|  | "[pharmacogenetics] can improve patient outcome as well. If you can treat the patient better, you know, it just gives your reputation, your clinic profile etc, [...] patients are happier." [GT11] (non-participating PCP) |
|  | "I don't think the patient would be willing to do this extra step. Because all they wanted to do is to try the medicine for about a few weeks [trial and error]" [GN03] (participating PCP) |
|  | "...more medical legal problem, right? If the test gives you a wrong answer and, uh, the patient suffers some adverse reaction, I think that is quite disastrous" [GT05] (non-participating PCP) |
|  | "I think software [recommendations] always reflect something. [...] The danger would be if you rely on your software, and the software is wrong" [GN01] (participating PCP) |
|  | "I cannot just change [the medication] because this [pharmacogenetic results] tells me to change, right? If they're happy with it and there's no adverse event [with the current treatment] and their blood pressure is under control [...] there's no reason for me to change" [GN02] (participating PCP) |
|  | "I've been taking Omeprazole and okay [Patient received results indicating potential allergy for Omeprazole]" [PN15] (participating-patient) |
|  | "Start this as a basic, then, [...] there could be a lot of trial and errors to get those library built up for a person, right?" [PT07] (non-participating patient) |
| 2. Appropriate financial context for implementation | “...without the Government subsidy, yeah, I don’t think people like we can afford it” [PN14] (participating patient) |
| | “...[to subsidise to] maybe below \$100” [PN08] |
| | “...if it’s going to be more than few \$100, then yeah, [...], the government [...] should subsidise, \$30 to \$50 would be a reasonable charge.” [PT07] (non-participating patient) |
|  | “It really depends on the amount that is going to be charged to the patient. [...] if he or she has been on a drug for a long time and they feel that there’s nothing wrong, [...] they may wonder why they are doing it. From a very practical perspective” [GN10] (participating PCP) |
|  | “I believe there are cost-effectiveness studies on this modality of, uh, testing, and [if] it proved to be of great value [...] then of course we will consider” “technology is there, it is scalable, the cost comes down with economy of scale, then I think, why not?” [GT05] (non-participating PCP) |
|  | “Private sector it’s all about, you know, cost-benefit ratio. If [...] it takes so much money to buy that kind of system [for pharmacogenetics] the return on investment has to justify in the long run [GT11] (non-participating PCP) |
| 3. Physician education and training in communicating results | “...If we get into my medical school days, you know, they, they talk about genetic testing during oncology posting” [GT11] (non-participating PCP) |
|  | “I’m the one who actually needs to explain the report to the patient. So if I don’t know how to explain it... I don’t know who I can consult” [GN03] (participating PCP) |
|  | “There’s actually about five, six, or seven [genes] only, right?... why they pick these special few, I mean, this maybe is not communicated to us also” [GN02] (participating-PCP) |
|  | “...they’ll do that [training] before ... But, then again, you can’t actually play with the software in reality [...] They don’t give you a test example” [GN01] (participating PCP) |
|  | “A point-of-contact will be very useful in case we see a report that we, um, that has alarm bells, and we just want to quickly, uh, address it” [GN10] (participating PCP) |
|  | “We don’t have actually a [link] directly with a geneticist, [...]or someone in the pharmacy industry, that we can rely on [...] to get some, er, help” [GN03] (participating PCP) |
|  | “I think it takes too much effort to explain all of these things to patients. [It] will be a huge hassle.” [GT11] (non-participating PCP) |
|  | "You say there's 'reduced efficacy.' What do you mean?, [...] there's increased adverse event? [...] What are the adverse events? These are all not available, so it's very difficult to [...] communicate this to the patients." [GN02] (participating PCP) |
|  | "The role of the doctor is important, how the doctor actually brings [...] the test to the attention of the patient, how it will benefit them." [GT05] (non-participating PCP) |
|  | "The issue is about patient education and patient literacy [...]. We need to do a lot in this respect." [GT04] |
|  | "Some of them don't understand why this genetic testing is necessary in the first place" [GN03] (participating PCP). |
|  | "...as long as you have the good therapeutic alliance with the patients, they're willing to listen to you" [GN19] (participating PCP) |
|  | "...again this is back to the shared decision-making process, [...], communicate with the patients, understand their values, and then [...] our suggestions, and see [...] which options did they choose." [GT05] (non-participating PCP) |
|  | "Ultimately, we are the clinicians, we make [] co-decision with the patient. So, if the information is presented and the patient is aware [...] patient says [...] I'm happy with what I'm on now..." [GN10] (participating PCP) |
|  | "I tend to trust them [doctors] when they pay attention to me, and then they spend quality amount of time. that's how's the rapport between me and the doctor is built." [GT05] (non-participating PCP) |
| 4. Integrating pharmacogenetic data into clinical records | "Yeah. It's very difficult to use. [...] We have to set up a different, [...] log in and then wait to, load, and then when you are looking at things, you, you cannot engage the patient, so that's not very good." [GN02] (participating PCP) |
|  | "Some people find it very naggy... they don't want any slowdown in their work"[GT04] (non-participating PCP) |
|  | "You have to go back two months to see that they were on this [pharmacogenetics] program. And sometimes you forget [...]. You are busy [...] It's the follow up that's not so easy... The initiation is easy" [GN19] (participating-PCP) |
|  | "The drugs are used in Canada, um, some of the drugs we use here are not reflected in the database." [GN03] (participating PCP) |
| 5. Data privacy and insurance implications | "Private information, so, I think it's, I guess, only natural that we have to take consent" [GT11] (non-participating PCP) |
|  | "They'll want to acquire this information. And then they will prequalify certain people, [...], and adjust their premium..." [PN13] (participating patient) |
|  | "...there will certainly be a few cases where people are denied [...] insurance based on a genetic predisposition." [GN01] (participating-PCP) |
|  | "So far all the patients didn't raise this issue about [their information] being exploited...I think it didn't occur to them." [GN03] (participating PCP) |
|  | "Yes, insurance is a problem. Employment... I don't think it's a problem. Most of the places you do not have to declare [ So, employment is based on whether you can perform the job [rather than health conditions]" [PT17] (non-participating patient) |

Five major themes were identified, 1. Perceptions of acceptability and test accuracy, 2. Appropriate financial context for implementation, 3. Physician education and training in communicating results, 4. Integrating pharmacogenetic data into clinical records, and 5. Data privacy and insurance implications.

### 1. Perceptions of acceptability and test accuracy

Professionals widely supported the concept of pharmacogenetic testing to inform prescribing, citing its potential to enhance patient safety and reduce adverse drug reactions. They recognized the potential to change the way prescribing decisions could be made, shifting from a ‘trial and error’ or ‘wait and see’ approach to an informed and patient-centred approach.

> *“We treat patients in a trial-and-error kind of way, […] pharmacogenetics reduces the amount of trial and error that you can have.” [GN19] (participating PCP)*

Patients described the potential benefits of their doctor being able to combine their traditional skills with pharmacogenetic testing and elect the optimal treatment with support from a clinical decision support system.

One clinician saw the benefits of pharmacogenetics extending beyond patient’s wellbeing, to enhancing the clinic’s reputation, saying *“[Pharmacogenetics] can improve patient outcomes as well. If you can treat the patient better, you know, it just gives your reputation, your clinic profile etc., […] patients are happier.” [GT11] (non-participating-PCP)*

Overall, the attitudes expressed were largely positive. The few comments expressing reluctance toward pharmacogenetic testing in primary care came from PCPs who were concerned about patients’ willingness, for example, *“I don’t think the patient would be willing to do this extra step. Because all they wanted to do is to try the medicine for about a few weeks [trial and error]” [GN03] (participating-PCP)*.

Despite broad support for pharmacogenetics, concerns about accuracy persisted. A few PCPs, regardless of experience, and several patients questioned the accuracy of pharmacogenetic results and clinical decision support recommendations. They feared errors could lead to inappropriate prescribing and medico-legal risks.

> *“…more medical legal problem, right? If the test gives you a wrong answer and, uh, the patient suffers some adverse reaction, I think that is quite disastrous” [GT05] (non-participating-PCP)*

Participating PCPs described unsettling scenarios where pharmacogenetic testing indicated a risk of adverse reactions in patients who had been taking medication for months without side effects and were deriving therapeutic benefit:

> *“I cannot just change [the medication] because this [pharmacogenetic results] tells me to change, right? If they’re happy with it and there’s no adverse event and their blood pressure is under control […] there’s no reason for me to change” [GN02] (participating-PCP)*.

### 2. Appropriate financial context for implementation

Both patients and doctors stressed that implementation of pharmacogenetics in primary care would require economic arrangements reflecting the price sensitive nature of Singaporean healthcare. Participants noted that the current high costs of genetic testing create a need for government subsidy to reduce out-of-pocket expenses.

> *“…without the Government subsidy, yeah, I don’t think people like we can afford it” [PN14] (participating-patient)*
>
> “Some patients shared ideas about the magnitude of subsidy that would be required, with comments such as *“$30 to $50 would be a reasonable charge.” [PT07] (non-participating-patient) or “below $100” [PN08] (participating-patient)*

Many drugs are first prescribed in community or primary care settings, making pre-emptive pharmacogenetic testing valuable by ensuring results are already in the patient’s record. However, in these settings, it may be challenging to convince patients of the perceived benefits, with a private PCP noting that decisions are shaped by cost-benefit considerations, return on investment, and business impact:

> *“Private sector it’s all about, you know, cost-benefit ratio. If […] it takes so much money to buy that kind of system [for pharmacogenetics] the return on investment has to justify in the long run [GT11] (non-participating-PCP)*.

### 3. Physician education and training in communicating results

Most of the PCPs discussed their personal sense of inadequate training in pharmacogenetics. Some recalled relevant teaching from oncology or genetic attachments during their training, *“…If we get into my medical school days, you know, they, they talk about genetic testing during oncology posting” [GT11] (non-participating-PCP).* For others, their knowledge came from self-directed learning from scientific literature.

PCPs who participated in the feasibility study received preparatory briefings and practice with a dummy case. While they found the training ‘adequate,’ they emphasized the need for further instruction on gene-specific testing indications and use of the interpretative software. Several also noted feeling isolated in practice and stressed the importance of having access to a pharmacogenetic expert for guidance when uncertainties arise.

> *“A point-of-contact will be very useful in case we see a report that we, um, that has alarm bells, and we just want to quickly, uh, address it” [GN10] (participating-PCP)*.

Whilst PCPs found their pre-test discussions with patients were largely without any challenges, they experienced greater difficulties communicating pharmacogenetic results and their relevance to prescribing decisions. Pharmacogenetic recommendations often indicate risk rather than certainty, and hence the interpretation and sharing of results with patients is more nuanced compared to routine and simple test results.

> *“You say there’s ‘reduced efficacy.’ What do you mean? […] there’s increased adverse event? […] What are the adverse events? These are all not available, so it’s very difficult to […] communicate this to the patients.” [GN02] (participating-PCP)*

The need for better lay understanding of pharmacogenetics was expressed repeatedly, including the rationale for testing, interpretation of the test results, and the immediate and long-term significance of findings for the individual.

> *“The issue is about patient education and patient literacy […]. We need to do a lot in this respect.” [GT04] (non-participating-PCP)*

Moreover, patients expressed a willingness to trust their doctors on pharmacogenetic matters, particularly if they were attentive and generous with their time:

> *“I tend to trust them [doctors] when they pay attention to me, and then they spend quality amount of time. that’s how’s the rapport between me and the doctor is built.” [PT17] (non-participating-patient)*.

### 4. Integrating pharmacogenetic data into clinical records

In the feasibility study, pharmacogenetic results were delivered through standalone software rather than being integrated into the electronic medical record. This created frustration, as clinicians had to access a separate system, adding time and disruption that was unsuitable for the high-volume patient throughput of primary care.

> *“Yeah. It’s very difficult to use. […] We have to set up a different [program], log in and then wait to load, and then when you are looking at things, you, you cannot engage the patient, so that’s not very good.” [GN02] (participating-PCP)*.

Participants highlighted the need for better Integration and tailoring of information to the local context as some commonly used medications in Singapore were not listed.

> *“The drugs are used in Canada, […], some of the drugs we use here are not reflected in the database.”[GN03] (participating-PCP)*.

### 5. Data privacy and insurance implications

Many participants stressed the need for formal consent and confidentiality of the pharmacogenetic information generated.

> *“Private information, so, I think it’s, I guess, only natural that we have to take consent” [GT11] (non-participating-PCP)*

All but one patient and all PCPs were apprehensive about pharmacogenetic information being shared with insurance companies and its potential negative impact on insurance premiums, either by refusing or withdrawing a policy or imposing higher premiums.

> *“They’ll want to acquire this information. And then they will prequalify certain people, […], and adjust their premium…”); [PN13] (participating-patient)*

Although discussions about health or life insurance were plentiful in our interviews, no clinician recalled a patient raising this as an issue in the feasibility study consultations.

> *“So far all the patients didn’t raise this issue about [their information] being exploited…I think it didn’t occur to them.” [GN03] (participating-PCP)*

## Discussion

This is the first study to explore patients and PCPs perceptions, attitudes, and beliefs about pharmacogenetic testing in Singapore. Discussions with 10 PCPs and 15 patients revealed broad support for pharmacogenetic testing, particularly in enhancing patient safety and reducing ADRs. Despite the apparent enthusiasm for pharmacogenetics there were multiple concerns identified that need addressing before implementation. These centred on test accuracy, the financial context for implementation, physician education and training in communicating results, integrating pharmacogenetic data into clinical records, and data privacy and insurance implications.

Singapore’s highly rated health care is organisationally complex, with public and private health services (17). Most residents use public facilities and are familiar with co-payments and government subsidies, and this explains why affordability plays a prominent role in patients’ expressed willingness to undergo pharmacogenetic testing (17). PCPs in the private sector were also aware that, however enthusiastic they were to adopt pharmacogenetic testing for their patients, without appropriate financial reimbursement, they would be unable to offer this service (18,19). In this context, Singapore may need to focus during implementation, as is happening in other countries, on the cost-effectiveness of pharmacogenetics from a societal perspective (15).

Among both participating and non-participating interviewees in the prior feasibility study, there was concern regarding confidentiality and the potential misuse of the pharmacogenetics data by the insurance industry. Singapore MOH’s Moratorium on Genetic Testing and Insurance prohibits insurers from mandating or encouraging individuals to undergo genetic testing as a prerequisite for insurance coverage (20). As yet there are no specific guidelines for pre-emptive or diagnostic pharmacogenetic testing, but the MOH has the experience of not dissimilar policy development.

PCPs recognised their need for enhanced education that would enable them to be effective as they moved beyond conducting the pharmacogenetic test to the correct interpretation of the results, giving meaningful explanations to their patients of the clinical utility of pharmacogenetics and how the findings impact the choice of medication for the patient. As well as the development of education resources and continuing medical education, some PCPs expressed a desire to have a helpline or mentor they could easily access if challenged with an unfamiliar or complex problem, an important consideration given that many private PCPs still work in solo practices.

Our study focused on providers and users of both public and private primary care but did not include other important stakeholders such as health administrators, policymakers, pharmacists, and healthcare professionals in other disciplines. These stakeholders may assist in identifying additional enablers to facilitate improved implementation of pharmacogenomics in Primary Care in the Singaporean context.

## Conclusions

Pharmacogenetic testing in primary care has the potential to significantly improve patient outcomes by reducing ADRs and optimizing drug choice and dosage. While both patients and PCPs expressed general support for pharmacogenetic testing in Singapore primary care, our study highlights several concerns about its successful integration and implementation, including cost, potential for misuse of genetic information by the insurance industry, and training adequacy. These concerns highlight the importance of considering financial incentives and subsidies, updating regulatory frameworks, and continuous education and engagement of stakeholders to enhance the feasibility and acceptability of pharmacogenetic testing in Singapore. Despite identified barriers, the favourable professional and patient attitudes and perceptions towards pharmacogenetic testing offer a fertile foundation for its gradual integration in primary care.

## Data Availability

All data produced in the present study are available upon reasonable request to the authors.

## Acknowledgments

Authors thank the patients and primary care physicians who participated in this study who were recruited through the Primary Care Research Network (pcRn), Singapore.

**Supplementary files 1: Semi-structured questionnaire used by the researchers to interview participating primary care physicians**

**Supplementary files 2: Semi-structured questionnaire used by the researchers to interview non-participating primary care physicians**

**Supplementary files 3: Semi-structured questionnaire used by the researchers to interview participating patients with chronic diseases**

**Supplementary files 4: Semi-structured questionnaire used by the researchers to interview non-participating patients with chronic diseases**

## Supplementary files 1: Semi-structured questionnaire used by the researchers to interview participating primary care physicians

**For the HCP taking part in the PGx study**

**Understanding the Professional and Patient Experiences or Concerns of Participation in Pharmacogenetics Testing in Primary Care in Singapore (PTPC)**

**Interview Guide**

**Introduction:**

*Introduce yourself and thank the interviewee for their participation*

During this interview we will discuss your views on pharmacogenetics, it will take approx. 30 to 40 minutes and will be digitally recorded. Your views will be anonymous for the transcription, analysis and publication. All information collected (including digital recording) will be stored securely, as per Nanyang Technological University policy, and destroyed after ten years.

**Review and sign the consent form, copy provided for the interviewee** ☐ Yes

**Confirm that the interview may be digitally recorded?** ☐ Yes / ☐ No

*Ask the interviewee if they have any questions or concerns*

**A. The Introduction**

These are some questions about you so that I can describe the HCP who helped with the research.

**Q1.** What type of Practice is yours and how many patients do you serve? How many patients do you see on a weekly basis? (private, group, polyclinic)

**B. The Core**

I would now like to explore your thoughts towards research in primary care

Perceptions

**Q2.** What is the role of research in your Practice? (other studies conducted at the Practice?)
**Q3.** What made you decide to take part in this study? (personal interest/ part of further training/ promotion)
**Q4.** What impact did this study have on your Practice? (time, workload)

I would now like to talk about your experience taking part in the PGx study

Attitudes

**Q5.** Did you have any anxieties about taking part in the study? (set up difficulties)
**Q6.** Any challenges/concerns with the consent process? (difficulties with explanation and interpretation)
**Q7.** What were the problems, if any, in practical aspects of taking the mouth swab and the de-identification process? (time, information, ease of execution)
**Q8.** Any challenges with the use of medical decision making system software? (was it easy to use/input information into/time to obtain results/ patient’s copy was this helpful)
**Q9.** Were the training and support sufficient to carry out the study? (recruitment/ equipment/guidance/communication with monitors)

Finally I would like to better understand your own beliefs in relation to PGx testing

Beliefs

**Q10.** What has changed in your medical Practice since taking part in the study? Probes
**Q11.** Would you be interested in other research related to PGx testing? Probes
**Q12.** What do you think will be the future for PGx testing in Singapore? (cost, implementation, patient attitudes, potential to reduce ADRs/any concerns re restrictions to prescribing/over reliance on MDSS/ do you agree with introducing evidenced based gene-drug algorithms)
**Q13.** What would you require in the future to undertake PGx testing? (support, financial incentive/setting for PGx primary care, secondary, tertiary, specialist dept)
**Q14.** Some people believe that racial or ethnic identity (e.g. Singapore Chinese, Singapore Indian, et.) can be used as a proxy for PGx testing result in predicting drug responses and thus PGx testing may not be needed. What do you think?

**Closing remarks**

Thank you for agreeing to speak to me today. Your comments have been very helpful.

Do you have any other questions or comments before we finish. Alternatively, please feel free to contact us later, our contact information is on your copy of the consent form.

## Supplementary files 2: Semi-structured questionnaire used by the researchers to interview non-participating primary care physicians

**For the HCP not taking part in the PGx study**

**Interview Guide**

**Introduction:**

*Introduce yourself and thank the interviewee for their participation*

**Review and sign the consent form, copy provided for the interviewee** ☐ Yes

**Confirm that the interview may be digitally recorded?** ☐ Yes / ☐ No

*Ask the interviewee if they have any questions or concerns*

**C. The Introduction**

These are some questions about you so that I can describe the HCPs who are interviewed for this study.

**Q1.** How many years have you been in practice? What type of practice is yours and how many patients do you serve? (private, group, polyclinic)

**D. The Core**

Awareness

**Q2:** What do you understand about pharmacogenetics? (fundamentals, applications, clinical utility)
**Q3:** Why do we need research studies in this area? (What are the unanswered questions?)
**Q4:** What is your experience in pharmacogenetic testing in your practice? (first-hand experience in primary care, experience in secondary/tertiary care, outcomes – positive/negative)

Perceptions

**Q5.** What are the research activities in your Practice? (studies conducted at the Practice)
**Q6.** Would you be interested if you were offered the opportunity to take part in a study utilising pharmacogenetic testing and a medical decision support software to improve prescribing for your patients? Why? (personal interest, part of further training/promotion, patient outcome)
**Q7.** What impact could the study have had on your Practice? (time, workload, patient outcomes)

Attitudes

**Q8.** Do you have any anxieties about inviting patients to participate or to consent to take part in a pharmacogenetic testing study? (set up difficulties, difficulties with explanation and interpretation)
**Q9.** Do you anticipate problems with the practical aspect of taking samples for genotyping and other research related activities? (time, information, ease of execution, de-identification process)
**Q10.** What is your understanding of medical decision support systems in healthcare? Have you ever previously used a medical decision support software? (was it easy to use/input information into, time to obtain results)
**Q11.** What additional information on the study that would be helpful to encourage GPs to take part in the study? What support would be helpful to encourage GPs to take part in the study? (guidance, communication, outreach, support, network)
**Q12.** Is there anything else that would affect your decision to take part in a PGx study? (thought patients would refuse, relevant local governance, insurance, cost)

Finally I would like to understand your own beliefs in relation to PGx testing

Beliefs

**Q13.** What do you think will be the future for PGx testing in Singapore? (cost, implementation, patient attitudes, role in potentially reducing ADRs, any concerns re restrictions to prescribing/over reliance on MDSS/ do you agree with introducing evidenced based gene-drug algorithms)
**Q14.** What would you require in the future to undertake research related to PGx testing? (support, financial incentive, setting for PGx in primary care, secondary, tertiary, specialist dept?)
**Q15.** Some people believe that racial or ethnic identity (e.g. Singapore Chinese, Singapore Indian, et.) can be used, as a proxy for PGx testing result, in predicting drug responses. Therefore, the need for PGx testing may be reduced. What do you think?

**Closing remarks**

Thank you for agreeing to speak to me today. Your comments have been very helpful.

## Supplementary files 3: Semi-structured questionnaire used by the researchers to interview participating patients with chronic diseases

**For the Patient taking part in the PGx study**

**<u>Interview Guide</u>**

*Introduce yourself and thank the interviewee for their participation*

**Review and sign the consent form, copy provided for the interviewee?** ☐ Yes

**Confirm that the interview may be digitally recorded?** ☐ Yes / ☐ No

*Ask the interviewee if they have any questions or concerns*

**A. The Introduction**

Before we go to the main topic for this interview I would like to get to know about you a bit better so I can put your thoughts in context

**Q1.** How long have you known your GP? (patient at this Practice, do you always see the same GP, good relationship)
**Q2.** How often do you see your GP? (only for repeat prescriptions/ more than 4 times a year, / are there any HCP that you visit more often/trust the most)

**B. The Core**

I would like to ask you some questions about what you thought about taking part in this research study

Perceptions

**Q3.** What motivated you to take part in the study? (did you discuss the study with anyone else / what appealed about the study)
**Q4.** What did your GP explain to you about the study? (explain the study to a friend what would you say/anything else you think relevant to the study that was not explained to you)
**Q5.** What did you expect to get out of this from participating in this study? (personalised care)
**Q6.** Have you heard about pharmacogenetics before? (friends/media/ had PGx testing before)
**Q7.** Did you find any challenge to your participation? If so, what challenge(s)? Is there anything we could have done to facilitate the process? (increased consultation time)
**Q9.** Did you have any anxiety about taking part in the study? If so, what anxieties? Is there anything we could have done to facilitate the research process?
**Q10.** How has your relationship to your GP changed since you participated? (improved/better communication/no change)

Now I would like to ask you about your current medication and the results you received from the study

Attitudes

**Q6.** Have you ever had a bad reaction from a prescribed medicine? (share experience/ was the medication stopped/changed/ were you hospitalised)
**Q7.** Did your medication change after receiving your PGx result from this study? Did you have a discussion with your GP about your results? (what did you learn/was there anything else you would have liked explained/ did you accept the change in medication/ if not why not/ any effects if you changed your medication/ any other changes to your care)
**Q8.** How do you think your results might be useful in the future? (do you have a copy of your results)

I would like to ask you about your thoughts on pharmacogenetics

Beliefs

**Q9.** Other than medication, do you think the PGx results will affect other areas of your care/life? (precision medicine/insurance/employment)
**Q10** Do you have any concerns regarding PGx? If so what are these? What could be done to ease your concerns? (reliability/time to receive results/ more medication prescribed as a result of test/more expensive medication/ lack of a suitable drug)
**Q11** Did you receive sufficient information on pharmacogenetics testing prior to agreeing to take part in the study? (did you have enough time to discuss the study/ask questions)
**Q12.** Based on your experience in the study, what do you think the challenges will be to implement this test in the Singapore population? What could be done to overcome these challenges? (confidentiality, government subsidies, treatment delivery and healthcare setting, incorporate as a part of routine practice)
**Q13.** Would you pay to have a pharmacogenetics test? (what would be a reasonable charge)

**C. Closing remarks**

Thank you for agreeing to speak to me today. Your comments have been very helpful.

## Supplementary files 4: Semi-structured questionnaire used by the researchers to interview non-participating patients with chronic diseases

**For the Patient not taking part in the PGx study**

**<u>Interview Guide</u>**

*Introduce yourself and thank the interviewee for their participation*

**Review and sign the consent form, copy provided for the interviewee?** ☐ Yes

**Confirm that the interview may be digitally recorded?** ☐ Yes / ☐ No

*Ask the interviewee if they have any questions or concerns*

**D. The Introduction**

**Q1.** How long have you known your GP? (patient at this Practice, do you always see the same GP, good relationship)
**Q2.** How often do you see your GP? (only for repeat prescriptions/ more than 4 times a year, are there any HCPs that you visit more often/trust more)

**E. The Core**

I would like to ask you some questions about what you thought about when discussing this research study

Perceptions

**Q3.** What could be done next time for you to participate? What would have to change? (did you discuss the study with anyone else/ any anxieties about the study/ what did not appeal about the study/ would you take part in other types of research)
**Q4.** What did your GP explain to you about the study? (explain the study to a friend what would you say/ anything else you think relevant to the study that was not explained to you?)
**Q5.** Have you heard about pharmacogenetics before? (friends/media/ had PGx testing before)
**Q6.** Have you participated in other health-related research before? What was different then?

Now I would like to ask you about your current medication

Attitudes

**Q7.** Have you ever had a bad reaction from a prescribed medicine? (share experience/ was the medication stopped/changed/ were you hospitalised/ does your medication change frequently)
**Q8.** How might PGx results change the treatment you receive? Be prepared to give a brief explanation of PGx in case they don’t know about it (medication/ precision medicine/ more medication/ more expensive)

I would like to ask you about your thoughts on pharmacogenetics

Beliefs

**Q9.** Other than medication, do you think the PGx results could affect other areas of your care/life? (precision medicine/insurance/employment)
**Q10** What are your concerns regarding PGx? (reliability/time to receive results/ more medication prescribed as a result of test/more expensive medication/ lack of a suitable drug)
**Q11** Did you receive sufficient information on pharmacogenetics testing prior to agreeing to take part in the study? (did you have enough time to discuss the study/ask questions)
**Q12.** Based on your experience, what do you think the challenges will be to implement this test in the Singapore population? What could be done to overcome these challenges? (confidentiality, government subsidies, treatment delivery, healthcare setting, incorporate as a part of routine practice?)
**Q13.** Would you pay to have a pharmacogenetics test? (what would be a reasonable charge)

**F. Closing remarks**

Thank you for agreeing to speak to me today. Your comments have been very helpful.

